# Understanding the association between immune modulating helminths and human papillomavirus or cervical cancer: a scoping review

**DOI:** 10.1101/2025.08.05.25330736

**Authors:** Allison Frank, Margarita Correa-Méndez, Alicia A. Livinski, Kalina Duncan, Eva H Clark, Patti Gravitt

## Abstract

**Background:** Cervical cancer disproportionately affects people living in low- and middle-income countries (LMICs). While access to effective preventive interventions explains some of the disparity, biological causes cannot be ruled out as important contributors to the observed disparities in cervical cancer outcomes. Because chronic infection with helminths which cause immune dysregulation and suppress anti-viral responses is common in LMICs, we sought to review the available epidemiologic evidence evaluating associations between helminth infection and HPV infection and/or cervical cancer.

**Methods:** We searched five databases of scientific publications using search terms targeted towards journal articles published between 1990-2022 evaluating an epidemiologic association between helminth infection and HPV prevalence, persistence, or cervical cancer progression. We identified eight relevant studies and describe them in this scoping review.

**Results:** All eight studies showed a positive population-level relationship between helminth infection and HPV or cervical cancer. Six studies found a positive association between schistosomiasis and cervical neoplasia; two studies found a positive association between hookworm infection and HPV prevalence; and 1 study found a positive association between *Ascaris*, *Trichuris*, and *Strongyloides* and HPV prevalence.

**Conclusions:** These data suggest a positive association between helminth infection and cervical neoplasia. Given the high burden of helminth and HPV co-infection in LMICs, further evaluation of helminth infection as a potential risk factor for cervical cancer development is warranted.

**Author Summary:** The high prevalence of helminth and HPV co-infections in LMICs significantly impacts quality of life, necessitating studies to expand our knowledge of their epidemiological and health impacts. Understanding the interaction between helminth infection, persistence of high-risk HPV, and cervical cancer is then crucial for developing evidence-based public health programs to improve health outcomes in affected populations. Chronic helminth infections, such as schistosomiasis and hookworm, induce immune responses that can impair antiviral responses critical for HPV clearance. Eight studies included in this scoping review suggest a positive association between helminth infection and cervical pre-cancerous and cancerous lesions, warranting further evaluation of this interaction given the high burden of co-infection. Findings expand the evidence-base to support the need to incorporate schistosomiasis screening into gynecological examinations and/or cervical cancer screening efforts to enhance the accuracy of diagnosing and treating pre- and cancerous cervical lesions in endemic regions.

## Introduction

In 2020, globally, over 604,000 people were diagnosed with cervical cancer and over 341,000 deaths were attributed to the disease (1). Although cervical cancer afflicts people across the globe, more than 85% of deaths occur in low- and middle-income countries (LMICs) (2).

Persistent infection with oncogenic human papillomavirus (HPV) types has been directly linked to cervical cancer development (3). While high-risk HPV (hrHPV) become undetectable within 6-18 months in more than 90% of infected individuals, hrHPV infections which remain persistently detectable have been shown to significantly increase the risk of progression to cancer (4–7). Well-established risk factors associated with hrHPV persistent detection and cancer development include HPV type, immune status, multiparity, and tobacco use (3, 8, 9). Cervical cancer rates vary significantly in LMICs where access to HPV vaccine and screening remains limited. While high HPV prevalence in the teens and early twenties likely reflects initiation of sexual activity, surveillance in many LMICs shows a second HPV prevalence peak in older women that is unrelated to sexual activity, or a relatively stable prevalence at all ages in some countries in sub-Saharan Africa (10). One hypothesis for what could be causing higher HPV prevalence in older women in LMICs is loss of immunologic control of HPV infection caused by an immune dysregulating agent. As well-demonstrated by HIV-HPV co-infection and high/early cervical cancer incidence in people living with HIV (PLWH), any agent that reduces the host’s ability to produce a robust Th1 (anti-viral) immune response during early HPV infection theoretically increases the host’s risk of persistent HPV infection and cervical cancer. Every effort should be made to identify and eliminate risk factors that permit hrHPV persistence to decrease cervical cancer risk.

Given the geographic overlap between LMICs with high cervical cancer rates and LMICs with high helminth burdens (Figures 1a and 1b), it has been hypothesized that the systemic immune dysregulation, as seen with HIV, can also be caused by chronic helminth infection (specifically schistosomiasis/bilharzia, hookworm, *Ascaris,* and *Trichuris*). Even a modest systemic immune dysregulation may facilitate hrHPV persistence, and thereby increase cervical cancer risk (8, 11–14). Further, chronic helminth infection may trigger direct and indirect changes to local immune microenvironments such as the cervico-vaginal mucosal immune system, changes that could impede hrHPV clearance (11). Urinary schistosomiasis also may act directly on the cervical epithelia (e.g., local inflammation from schistosomiasis eggs) to make it easier for cervical cancer to develop (12). Any co-infection which (a) allows HPV to alter the host’s immune response and/or (b) increases local inflammation potentially increases the risk of developing cervical cancer for people with HPV and may lead to earlier onset disease.

**Fig 1:**
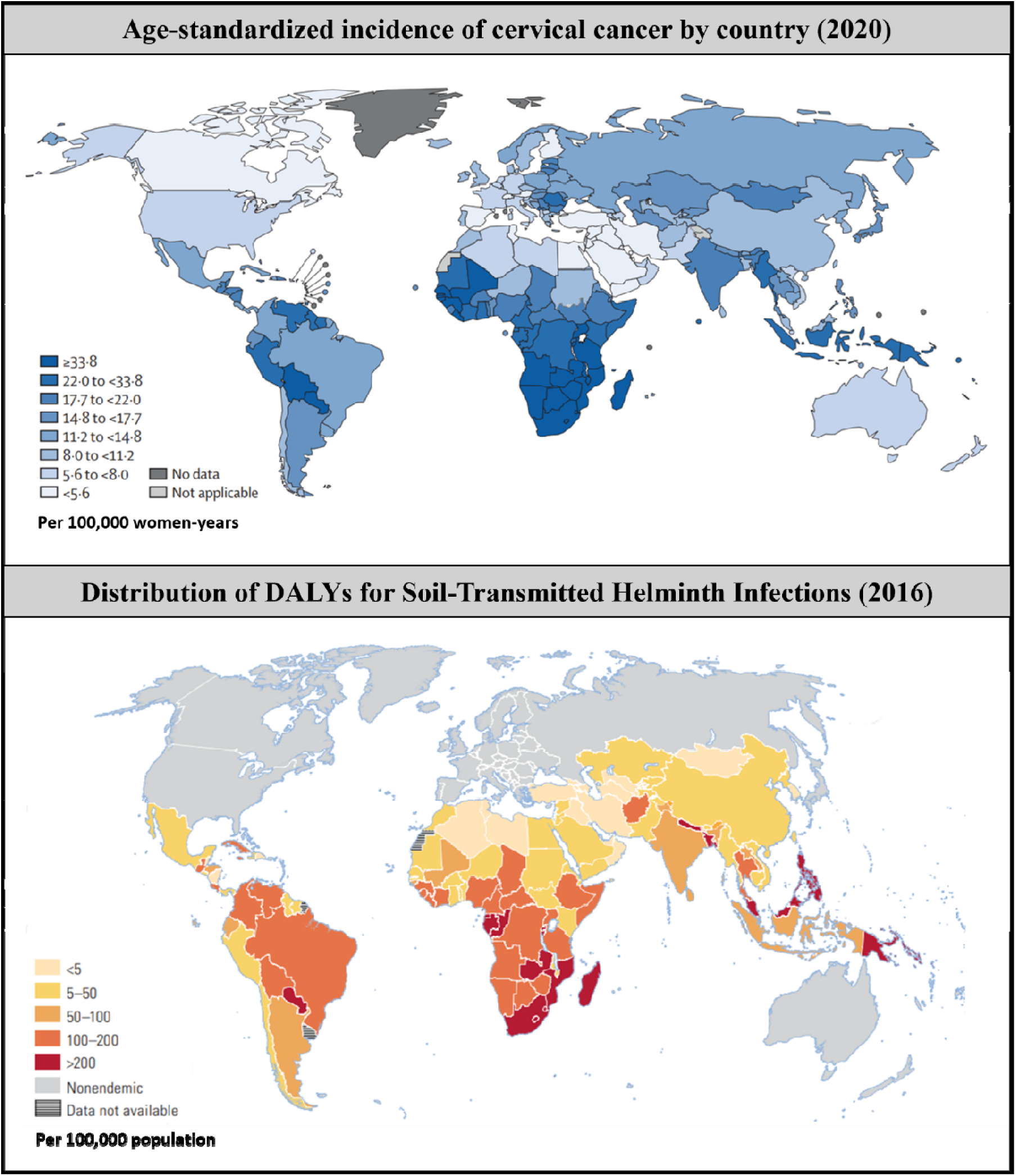
Geographic distribution of cervical cancer incidence rates and helminth burden A. Age standardized incidence rates of cervical cancer in 2020 (13, 15) B. Distribution of disability-adjusted life years (DALYs) for Soil-Transmitted Helminth Infections (hookworm, roundworm, and whipworm), per 100,000 Population (14)

This scoping review focuses specifically on two research questions: (1) what epidemiological evidence exists describing an association between helminth infection and HPV prevalence, persistence, or progression of dysplasia and (2) what gaps exist in the research? Although basic biology related to this topic has been reviewed elsewhere, this scoping review will summarize epidemiologic research in this area (11). The findings of this review will further our understanding on helminth infections as risk factors for HPV infections and cervical cancer, providing helpful insights to better inform and strengthen public health programs that seek to facilitate access to deworming treatments and cervical cancer prevention and control programs in LMICs.

## Methods

We followed the Joanna Briggs Institute (JBI) methodology and used the Preferred Reporting Items for Systematic reviews and Meta-Analyses extension for Scoping Reviews (PRISMA-ScR) Checklist for reporting (S1 PRISMA checklist) (16–18).

### Eligibility Criteria

We included population and clinical studies (e.g., cohort, case-control, follow-up, observational, retrospective, or prospective studies) conducted in LMICs that addressed helminth infections, including schistosomiasis/bilharzia, hookworms, *Ascaris*, and *Trichuris*, and either HPV or cervical cancer. We included original primary research articles, conference abstracts/proceedings, conference papers, books, and book chapters published between 1990 and 2022. We selected 1990 as the cutoff date since this was when high quality HPV (PCR-based) testing was introduced. We included manuscripts written in English or Spanish language. We excluded studies conducted in the United States or other upper-middle or high-income countries because helminth infections are not endemic in these countries. We also excluded studies that did not specify the type of helminth studied, reported on animal- or cell-based laboratory studies, or focused on assessing the association between helminth infections and other types of cancers (e.g., bladder cancer). We excluded systematic reviews and meta-analysis, but we scanned the references of relevant reviews to identify other potentially relevant articles. We excluded editorials, letters, and case studies.

### Information Sources

We defined our search strategy in collaboration with an NIH biomedical librarian (AAL), who conducted the database searches. We conducted the search for studies published after 1990 using five databases: Embase (Elsevier); Global Health (CABI Abstracts); PubMed (National Library of Medicine); Scopus (Elsevier); and Web of Science: Core Collection (Clarivate Analytics).

### Search

Search terms included keywords and controlled vocabulary terms (e.g., MeSH, EMTREE) for each concept of interest (helminths, HPV, cervical cancer, LMICs) (see Supplemental file 2 for full list of search terms for each database). EndNote 20 (Clarivate Analytics) was used to collect, manage, and identify duplicate records from the literature searches. Additionally, the reference lists of included articles and relevant systematic reviews and meta-analyses were scanned by two reviewers to identify other potentially relevant articles. Any articles identified in this manner were then screened using the procedure below.

### Selection of Sources of Evidence

Two reviewers (AF and MCM) piloted the screening process on a random sample of records selected by the biomedical librarian. Resulting changes to the screening process and eligibility criteria were completed prior to commencing the official screening steps. The reviewers independently screened each unique record retrieved from the literature searches via a two-step screening process using Covidence (Veritas Health Innovations). First, the reviewers screened all titles and abstracts using the established eligibility criteria. Second, records remaining after title and abstract review underwent full text screening using the same eligibility criteria. The reviewers met weekly to discuss disagreements and reach consensus. When needed, other members of the team informed screening decisions and resolved discrepancies.

### Data Charting Process

Once the final set of included records was identified, the two reviewers developed a data extraction codebook and piloted it using a randomly selected subset of 15 records. Both reviewers independently collected data using Covidence. Discrepancies between the two reviewers were resolved by a third author (PG).

### Variables

The variables extracted from each manuscript included: title, author, publication date, country and setting of study, study design and objectives, age range of participants, type of exposure studied (i.e., type of helminth), methods to measure exposure status, outcome reported, methods to measure outcome status, methods to measure how exposure and outcome related to one another, main analysis method, primary outcome as related to HPV or cervical cancer and helminth infection, as well as comments, limitations and gaps raised by the study authors. The data extraction codebook and additional details on data collection are available in the supplemental materials (Supplemental file 3).

### Synthesis of Results

Extracted data were organized by authors AF and PG, who grouped exposures into four categories: schistosomiasis/bilharzia, hookworms, *Ascaris*, and *Trichuris*. Since the outcome for this study (HPV or cervical cancer) was relatively broad, study outcomes were synthetized based on what was specifically reported in each study. Included outcomes were cervical cancer, cervical intraepithelial neoplasia (CIN), high grade squamous intraepithelial lesion (HSIL), and HPV prevalence.

Positive associations were determined based on positive effect measurements stated in the study or calculated by authors. For some studies, OR and 95% CIs were not directly reported in the manuscripts, but they were calculated by authors (AF and PG) based on the 2×2 tables that were derived directly from the data reported (Table 1).

**Table 1:**
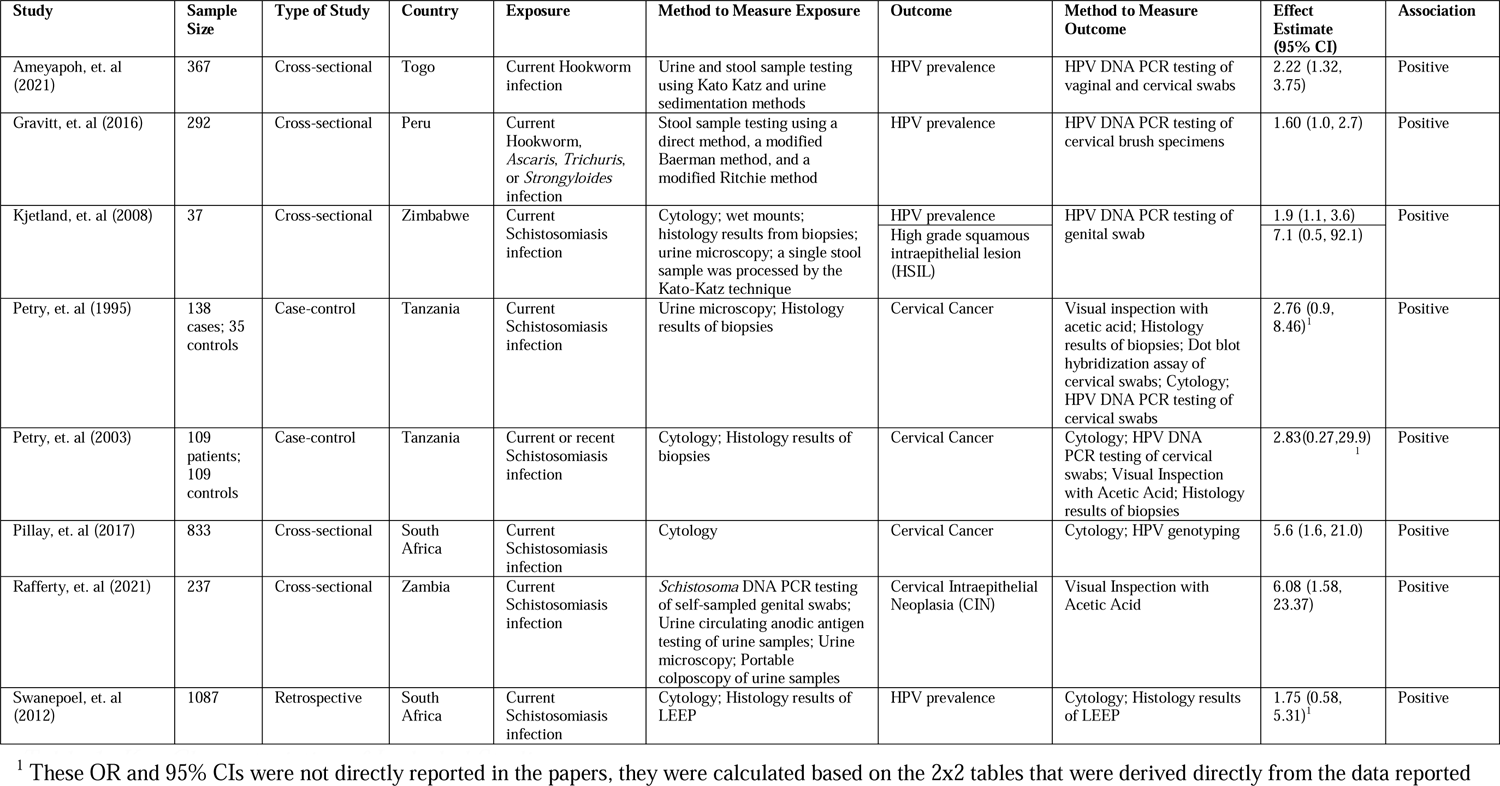
Key Characteristics of Included Studies.

## Results

### Selection of Sources of Evidence

As shown in Figure 2, 2309 articles were retrieved from the database searches, of which 1085 were duplicates. We screened the titles and abstracts of 1224 articles and, of those, excluded 1131 based on the eligibility criteria. Ninety-three articles that matched eligibility criteria in the title and abstract underwent full text. Eight underwent final data extraction and were included in this review. Of the 85 excluded, 31 were case studies, 12 did not explore helminth/HPV or cervical cancer association, 11 were editorials or letters, 11 were reviews/meta-analyses, eight contained inadequate information, five were not epidemiological studies, four discussed cancer other than cervical cancer, one was written before 1990, one did not specify helminth type, and one was in a language other than English or Spanish.

**Fig 2:**
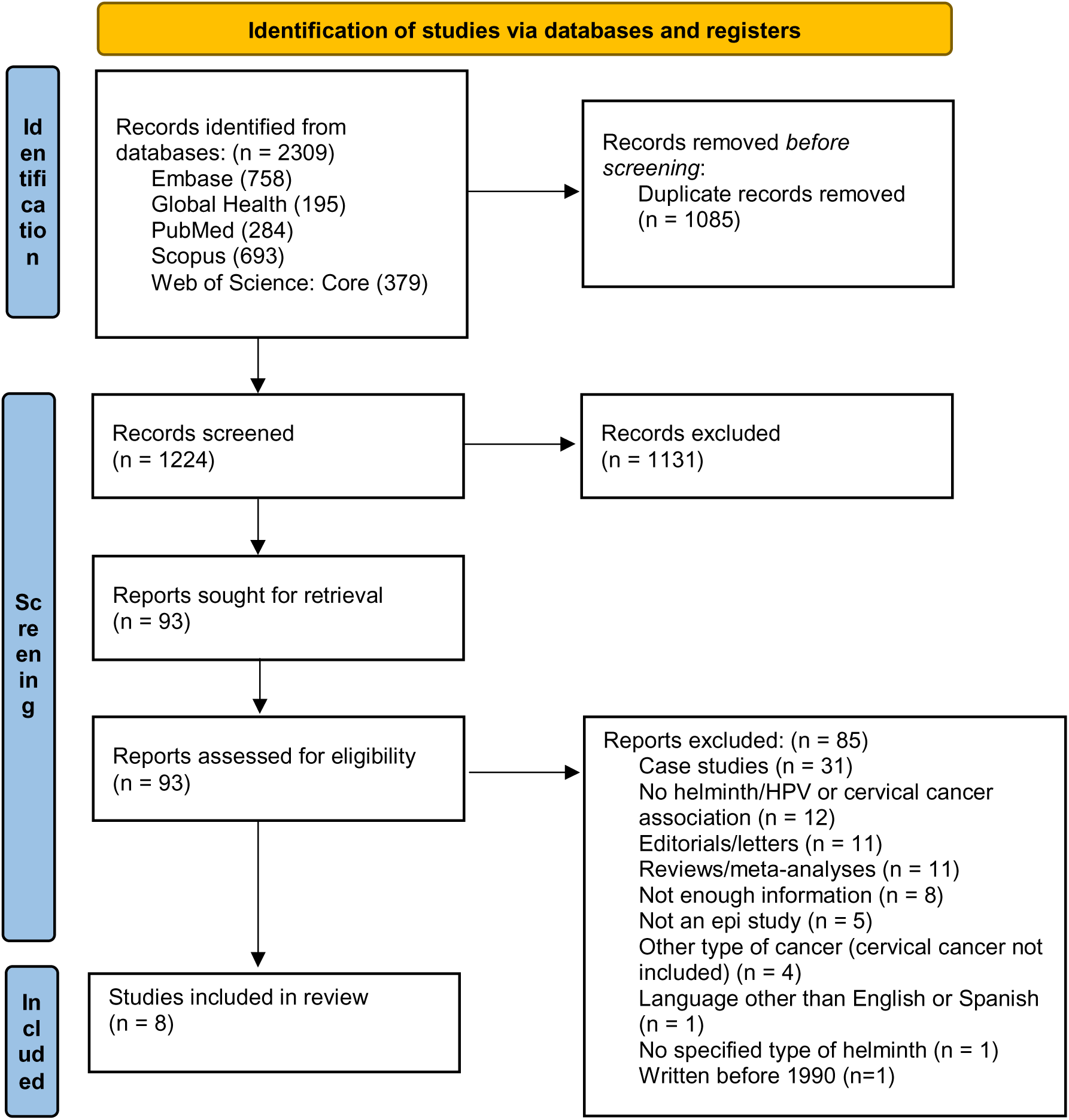
PRISMA Flow Diagram.

### Characteristics of Sources of Evidence

Of the eight studies included in this review, five studies were cross-sectional, two were case-control, and one was retrospective (Table 1). Seven of the eight studies were conducted in Africa, which included five countries. One study was conducted in Peru. See Table 1 for a full breakdown of data characteristics.

### Exposures

Regarding exposure, six studies evaluated participants for schistosomiasis (all in Africa) and two evaluated participants for intestinal helminth infections. Specimens used to evaluate participants for helminth infection varied by study (Table 1). The six studies that evaluated their participants for schistosomiasis did so by either cytology; histology results from loop electrosurgical excision procedure (LEEP) or biopsies; portable colposcopy of urine samples; stool samples processed by the Kato-Katz technique; Schistosoma DNA PCR testing of self-sampled genital swabs; urine circulating anodic antigen testing of urine samples; urine microscopy; or wet mounts. The two studies that evaluated their participants for just hookworm did so by either urine and stool sample testing using Kato Katz and urine sedimentation methods. The one study that evaluated their participants for Hookworm, *Ascaris, Trichuris,* or *Strongyloides* infection did so by stool sample testing using a direct method, a modified Baerman method and a modified Ritchie method. See Table 1 for a full breakdown of exposure measurement data.

### Outcomes

The measured outcome was HPV prevalence for four studies, cervical cancer for three, CIN for one, and HSIL for one. Methods to measure each type of outcome varied by study (Table 1). Of the four studies that evaluated HPV prevalence as the outcome, three used HPV DNA PCR testing and one used cytology and histology results of LEEP. Of the three studies that evaluated cervical cancer as the outcome, three used cytology; two used histology results of biopsies; two used HPV DNA PCR testing; two used visual inspection with acetic acid (VIA); one used dot blot hybridization assay of cervical swabs; and one study conducted HPV genotyping. See Table 1 for a full breakdown of outcome measurement data.

### Epidemiologic associations between helminths and hrHPV-related cervical dysplasia or cancer

#### Schistosomiasis and hrHPV-related cervical dysplasia/cancer

All six studies that explored the relationship between schistosomiasis and cervical neoplasia found a positive association between the two conditions. Kjetland et al’s study in Zimbabwe demonstrated that women with schistosomiasis were more likely to have hrHPV (OR: 1.9; 95% CI: 1.1–3.6) as well as HSIL (OR: 7.1; 95% CI: 0.5,92.1) (19). The Zambian study showed similarly high risk of CIN (OR: 6.08; 95% CI: 1.58–23.37) for women with schistosomiasis (20). Swanepoel et al’s retrospective study of South African women with HIV reported that participants with schistosomiasis had 75% higher odds of CIN2+ or cervical cancer compared with women whose biopsies showed CIN1 or koilocytosis (OR: 1.75; 95% CI: 0.58–5.31) (21). Pillay et al’s South African study (OR: 5.6; 95% CI: 1.6–21.0) found a positive association between schistosomiasis and cervical cancer (22). Similarly, both Tanzanian studies, the Petry et al. study (1995) (OR: 2.76; 95% CI: 2.76–8.46) and the Petry et al study (2003) (OR: 2.83, 95% CI: 0.27–29.96) found a positive association between schistosomiasis and cervical cancer (23, 24).

#### Intestinal helminth infection and hrHPV-related cervical dysplasia/cancer

Gravitt et al’s Peruvian study demonstrated that women with intestinal helminth infection were more likely to be HPV positive (OR: 1.6; 95% CI: 1.0–2.7) (11). Ameyapoh et al.’s Togo study similarly showed a positive association between hookworm infection and cervical neoplasia (OR: 2.22; 95% CI: 1.32–3.75) (25).

## Discussion

The results of this study show consistent positive associations between helminth infections and either cervical HPV persistence or cervical precancerous dysplasia, but not invasive cervical cancer. Because there are positive associations between immunomodulatory helminth infections and cervical neoplasia but not cervical cancer, these epidemiologic data are consistent with helminths playing a role in the immune dysregulation of the oncogenic virus HPV instead of playing a role as a direct carcinogen for cervical cancer (e.g., as is the case for schistosomiasis and bladder cancer) (11). Since the data suggests a positive association between helminth infection and cervical intraepithelial neoplasia and given the high burden of helminth and HPV co-infection in LMICs, whose populations bear 85% of the global cervical cancer burden, further evaluation of this potential interaction is warranted. Not all effect measurements were statistically significant, however they did all trend towards an increased risk. This is likely due to the small sample sizes of most studies. Given the consistent direction of these association, additional studies are needed to broaden our understanding.

Although these studies do not provide evidence of helminth infection as a direct carcinogen independent of HPV, authors suggest the potential link between schistosomiasis infection and DNA damage which accelerates carcinogenesis of hrHPV, so this mechanism cannot be ruled out from these studies. Another hypothesis is that helminths work upstream in the causal pathway by increasing risk of persistent HPV detection by activating the Th2 immune response, which in turn downregulates the Th1 response needed to control viral infections like HPV (8). Some hypothesize that schistosomiasis can lead to HPV persistence through damage to the cervical epithelium and an additional DNA damage inflammation carcinogenic effect (26). Additionally, schistosomiasis could cause local immune modulation, providing a tolerogenic environment for HPV to persist (26). While more research is needed to test these hypotheses, known possible mechanisms for the association give important biological credibility to the findings.

This study has several limitations. Amongst the scope of this review, there is a likelihood of positive reporting bias, therefore there may be a lack of publications on studies that did not find an association. Additionally, not all studies adjusted for important confounding effects of sexual behaviors and other immune suppressive infections (e.g., HIV). To the extent that these exposures were differential between our helminth exposure groups, the point estimates may be biased. Since only papers written in English or Spanish language were included in this study, there is a chance important data from papers written in other languages was missed. Any data from studies published before 1990 that could be relevant to this topic was not looked at and furthermore, studies published in local journals and not indexed in the journals included in this review could have been missed. As the search for this scoping review was conducted in April 2022, we might have missed some recent publications or grey literature published since then.

### Public Health Impact

The World Health Organization’s road map to eliminate neglected tropical diseases (NTDs) by 2030 outlines strategies to prevent, control, and eliminate NTDs, including helminth infection (27). Strategies include improvements in water, sanitation, and hygiene (WASH), innovation in diagnostics, and increased access to deworming treatments (27). Recognizing the overlap between HrHPV and helminth infections, public health programs that bundle cervical cancer and helminth prevention and control programs could have significant impacts. Efforts to understand the interaction between helminth infection, persistence of hrHPV, and cervical cancer, are important to increase the evidence-base for these public health programs and improve health outcomes in the population (8, 11).

## Conclusion

Considering this high prevalence and the overlap of HPV and helminth infection, and the impact these infections have on the quality of life of populations in LMICs, studies that help expand our understanding on the epidemiological evidence and health impact of these infections are essential. The associations identified through this scoping review contribute towards the evidence-base that can be helpful to inform public health programs and guidelines. The results summarized here are also important to increase awareness of the need to consider Schistosomiasis screening in gynecological examinations to improve the accuracy in diagnosis and treatment of pre- and cancerous lesions. Since helminth re-infection is common in individuals living in endemic areas, there are steps that should be taken to prevent re-infection, including frequent (every three months) de-worming of reproductive age individuals and improvements to WASH. It is then important to consider the syndemics of helminths and HPV infections (and likely many other carcinogenic viral infections) and how they change the individual and population ecology, especially in rural areas where cervical cancer is high [(28). Given major public health initiatives for NTDs and HPV/cervical cancer, we have a time-sensitive opportunity for cross-disciplinary collaboration to more fully understand the relationships between these infections and their causal pathways as well as the biological mechanisms of interactions to enable the identification of effective public health interventions.

## Supporting information

Supplemental Information 1 - PRISMA ScR

Supplemental File 2 - Search Strategy and Databases Searched

## Data Availability

All data underlying the findings of this study are within the manuscript and its supporting information files

## References

1. Singh D, Vignat J, Lorenzoni V, Eslahi M, Ginsburg O, Lauby-Secretan B, et al. Global estimates of incidence and mortality of cervical cancer in 2020: a baseline analysis of the WHO Global Cervical Cancer Elimination Initiative. Lancet Glob Health. 2023;11(2):e197–e206.

2. Hull R, Mbele M, Makhafola T, Hicks C, Wang SM, Reis RM, et al. Cervical cancer in low and middle-income countries. Oncol Lett. 2020;20(3):2058–74.

3. Walboomers JM, Jacobs MV, Manos MM, Bosch FX, Kummer JA, Shah KV, et al. Human papillomavirus is a necessary cause of invasive cervical cancer worldwide. The Journal of pathology. 1999;189(1):12–9.

4. Plummer M, Schiffman M, Castle PE, Maucort-Boulch D, Wheeler CM, Group A. A 2-year prospective study of human papillomavirus persistence among women with a cytological diagnosis of atypical squamous cells of undetermined significance or low-grade squamous intraepithelial lesion. The Journal of infectious diseases. 2007;195(11):1582–9.

5. Rosa MI, Fachel JM, Rosa DD, Medeiros LR, Igansi CN, Bozzetti MC. Persistence and clearance of human papillomavirus infection: a prospective cohort study. Am J Obstet Gynecol. 2008;199(6):617 e1-7.

6. Miranda PM, Silva NN, Pitol BC, Silva ID, Lima-Filho JL, Carvalho RF, et al. Persistence or clearance of human papillomavirus infections in women in Ouro Preto, Brazil. BioMed research international. 2013;2013:578276.

7. Schiffman M, Castle PE, Jeronimo J, Rodriguez AC, Wacholder S. Human papillomavirus and cervical cancer. Lancet. 2007;370(9590):890–907.

8. Clark EH, Gilman RH, Chiao EY, Gravitt PE. Gut Helminth Infection-Induced Immunotolerance and Consequences for Human Papillomavirus Persistence. The American journal of tropical medicine and hygiene. 2021;105(3):573–83.

9. Muñoz N, Franceschi S, Bosetti C, Moreno V, Herrero R, Smith JS, et al. Role of parity and human papillomavirus in cervical cancer: the IARC multicentric case-control study. Lancet. 2002;359(9312):1093–101.

10. de Sanjosé S, Diaz M, Castellsagué X, Clifford G, Bruni L, Muñoz N, Bosch FX. Worldwide prevalence and genotype distribution of cervical human papillomavirus DNA in women with normal cytology: a meta-analysis. Lancet Infect Dis. 2007;7(7):453–9.

11. Gravitt PE, Marks M, Kosek M, Huang C, Cabrera L, Olortegui MP, et al. Soil-Transmitted Helminth Infections Are Associated With an Increase in Human Papillomavirus Prevalence and a T-Helper Type 2 Cytokine Signature in Cervical Fluids. The Journal of infectious diseases. 2016;213(5):723–30.

12. Sturt AS, Webb EL, Patterson C, Phiri CR, Mweene T, Kjetland EF, et al. Cervicovaginal Immune Activation in Zambian Women With Female Genital Schistosomiasis. Front Immunol. 2021;12:620657.

13. Cervix Uteri 2020. 2020.

14. Bundy DAP, Appleby Laura J, Bradley M, Croke K, Hollingsworth TD, Pullan R, et al. Mass Deworming Programs in Middle Childhood and Adolescence. In: Bundy DAP, Silva ND, Horton S, Jamison DT, Patton GC, editors. Child and Adolescent Health and Development. 3rd ed. Washington (DC)2017.

15. Singh D, Vignat J, Lorenzoni V, Eslahi M, Ginsburg O, Lauby-Secretan B, et al. Global estimates of incidence and mortality of cervical cancer in 2020: a baseline analysis of the WHO Global Cervical Cancer Elimination Initiative. The Lancet Global Health. 2023;11(2):e197–e206.

16. Moher D, Shamseer L, Clarke M, Ghersi D, Liberati A, Petticrew M, et al. Preferred reporting items for systematic review and meta-analysis protocols (PRISMA-P) 2015 statement. Systematic reviews. 2015;4(1):1.

17. Kuper H, Adami HO, Trichopoulos D. Infections as a major preventable cause of human cancer. Journal of Internal Medicine. 2000;248(3):171–83.

18. Tricco AC, Lillie E, Zarin W, O’Brien KK, Colquhoun H, Levac D, et al. PRISMA Extension for Scoping Reviews (PRISMA-ScR): Checklist and Explanation. Annals of internal medicine. 2018;169(7):467–73.

19. Kjetland EF, Kurewa EN, Ndhlovu PD, Midzi N, Gwanzura L, Mason PR, et al. Female genital schistosomiasis - a differential diagnosis to sexually transmitted disease: genital itch and vaginal discharge as indicators of genital Schistosoma haematobium morbidity in a cross-sectional study in endemic rural Zimbabwe. Tropical Medicine & International Health. 2008;13(12):1509–17.

20. Rafferty H, Sturt AS, Phiri CR, Webb EL, Mudenda M, Mapani J, et al. Association between cervical dysplasia and female genital schistosomiasis diagnosed by genital PCR in Zambian women. BMC Infect Dis. 2021;21(1):691.

21. Swanepoel PJ, Michelow P, Du Plessis R, Proudfoot IG, Tarr GA, Bockel SL, Swanepoel CJ. Cervical squamous intraepithelial lesions and associated cervical infections in an HIV-positive population in Rural Mpumalanga, South Africa. Cytopathology. 2013;24(4):264–71.

22. Pillay P, Taylor M, Galappaththi-Arachchige H, Christiansen IK, Ambur OH, Roald B, Kjetland E. Liquid based cytology for diagnosis and risk assessment of cervical atypia in schistosoma and HIV endemic populations. Journal of lower genital tract disease. 2017;21(2):S24.

23. Petry KU, Kochel H, Kupsch E, Kingu H. The association of Schistosoma haematobium, human papillomavirus and cervical neoplasia in a rural setting in tropical East Africa. Cervix and the Lower Female Genital Tract. 1995;13(1):23–7.

24. Petry KU, Scholz U, Hollwitz B, Von Wasielewski R, Meijer CJ. Human papillomavirus, coinfection with Schistosoma hematobium, and cervical neoplasia in rural Tanzania. Int J Gynecol Cancer. 2003;13(4):505–9.

25. Holali Ameyapoh A, Katawa G, Ritter M, Tchopba CN, Tchadié PE, Arndts K, et al. Hookworm Infections and Sociodemographic Factors Associated With Female Reproductive Tract Infections in Rural Areas of the Central Region of Togo. Front Microbiol. 2021;12:738894.

26. Wu Y, Duffey M, Alex SE, Suarez-Reyes C, Clark EH, Weatherhead JE. The role of helminths in the development of non-communicable diseases. Front Immunol. 2022;13:941977.

27. Casulli A. New global targets for NTDs in the WHO roadmap 2021-2030. PLoS Negl Trop Dis. 2021;15(5):e0009373.

28. Hulme A, Thompson J, Brown A, Argus G. The need for a complex systems approach in rural health research. BMJ open. 2022;12(10):e064646.

