## Supplemental File 2 - Search Strategy and Databases Searched for "Understanding the association between immune modulating helminths and human papillomavirus or cervical cancer: a scoping review"

**Title of Review:** Understanding the association between immune modulating helminths and human papillomavirus or cervical cancer: a scoping review

**Database:** PubMed/MEDLINE
**Platform:** US National Library of Medicine
**Date Searched:** April 11, 2022

**Database Date Coverage:** 1946–present
**Date Limits:** Publication date: 1/1/1990–12/31/2022
**Other Limits/Filters:** Language: English & Spanish

| **Set** | **Concept** | **Search Strategy** |
| --- | --- | --- |
| #1 |  | (((HPV[Title/Abstract] OR “cervical cancer*”[Title/Abstract] OR “cervical neoplasm*”[Title/Abstract] OR “cervical neoplasia*”[Title/Abstract] OR "Uterine Cervical Neoplasms"[Mesh] OR "Cervical Intraepithelial Neoplasia"[Mesh] OR “cervical intraepithelial neoplasia*”[Title/Abstract] OR "Papillomaviridae"[Mesh:NoExp] OR "Papillomavirus Infections"[Mesh] OR "cervix tumour*"[Title/Abstract] OR "cervix tumor*"[Title/Abstract] OR "cervix cancer*"[Title/Abstract] OR "cervix carcinoma*"[Title/Abstract] OR "cervical tumor*" [Title/Abstract] OR "cervical tumour*" [Title/Abstract] OR papillomavirus*[Title/Abstract] OR “papilloma virus*”[Title/Abstract]) NOT (spinal[Title/Abstract] OR spine[Title/Abstract])) OR ((cancer[Title/Abstract] OR cancers[Title/Abstract] OR cancerous[Title/Abstract] OR neoplasm*[Title/Abstract] OR carcinogen*[Title/Abstract]) AND (cervix[Title/Abstract] OR cervical[Title/Abstract]))) |
| #2 |  | ("Parasitic Diseases"[Mesh:NoExp] OR parasite*[Title/Abstract] OR parasitic[Title/Abstract] OR parasitosis[Title/Abstract] OR helminth*[Title/Abstract] OR ectoparasit*[Title/Abstract] OR "Helminths"[Mesh:NoExp] OR "Helminthiasis"[Mesh] OR "Schistosoma"[Mesh] OR Schistosomiasis[Mesh] OR schistosom*[Title/Abstract] OR "Hookworm Infections"[Mesh] OR hookworm*[Title/Abstract] OR "hook worm*"[Title/Abstract] OR ancylostomatoid*[Title/Abstract] OR ancylostom*[Title/Abstract] OR bunostomum*[Title/Abstract] OR necator*[Title/Abstract] OR "Ancylostomatoidea"[Mesh] OR "Ancylostoma"[Mesh] OR "Necator"[Mesh] OR "[Trichuriasis"[Mesh](https://www.ncbi.nlm.nih.gov/mesh/68014257)] OR "Trichuris"[Mesh] OR trichuri*[Title/Abstract] OR whipworm*[Title/Abstract] OR “whip worm*”[Title/Abstract] OR bilharzia*[Title/Abstract] OR bilharziosis[Title/Abstract] OR ascari*[Title/Abstract] OR "Ascaris"[Mesh] OR "Ascariasis"[Mesh]) |
| #3 |  | #1 AND #2 |
| #4 |  | #3 AND (English[lang] OR Spanish[lang]) AND (("1990/01/01"[Date - Publication] : "2022/12/31"[Date - Publication])) |

**Notes:** The limits for publication year (1990–2022) and language (Spanish and English) were applied to the main search using the filters available in PubMed. The keywords were searched in the title and abstract fields in PubMed (i.e., [Title/Abstract]), and the controlled vocabulary terms are indicated with [Mesh] or if the MeSH term was not exploded to automatically include all narrower terms this was indicated with [Mesh:Noexp]. Phrases were enclosed in quotation marks to force the searching of the exact terms in order presented. No other limits were applied to the searches.

**Database:** Web of Science: Core Collection*
**Platform:** Clarivate Analytics
**Date Searched:** April 11, 2022

**Database Date Coverage:** 1900–present
**Date Limits:** Publication date: 1990–2022
**Other Limits/Filters:** Language: English & Spanish

| **Set** | **Concept** | **Search Strategy** |
| --- | --- | --- |
| #1 | HPV or Cervical Cancer | TS=((((HPV OR 'cervical cancer*' OR 'cervical neoplasm*' OR 'cervical neoplasia*' OR 'cervical intraepithelial neoplasia*' OR papillomavirus* OR 'papilloma virus*' OR 'cervix tumour*' OR 'cervix tumor*' OR 'cervix cancer*' OR 'cervix carcinoma*' OR 'cervical tumor*' OR 'cervical tumour*' OR 'uterine cervix tumor' OR 'uterine cervix cancer' OR 'uterine cervix carcinoma' OR 'uterine cervix carcinoma in situ' OR 'Wart virus' OR 'papillomavirus infection') NOT (spine OR spinal)) OR ((cancer OR cancers OR cancerous OR neoplasm* OR carcinogen*) AND (cervix OR cervical)))) |
| #2 | Helminths | TS=(parasite* OR parasitic OR parasitosis OR helminth* OR ectoparasit* OR schistosom* OR hookworm* OR 'hook worm*' OR ancylostomatoid* OR ancylostom* OR bunostomum* OR necator* OR trichuri* OR whipworm* OR 'whip worm*' OR bilharzia* OR bilharziosis OR ascari*)) |
| #3 |  | #1 AND #2 |
| #4 | Limits Applied | #3 Apply filters for language (English and Spanish) and publication year (1990–2022) |

**Notes:** The limits for publication year (1990–2023) and language (English and Spanish) were applied to the main search using the filters available. The keywords were searched in the Topic field (i.e., TS) which searches the title, abstract, author keywords, and KeyWords Plus fields. Phrases were enclosed in quotation marks to force the searching of the exact terms in order presented. No other limits were applied to the searches.

*Science Citation Index Expanded (SCI-EXPANDED)--1900-present

Social Sciences Citation Index (SSCI)--1900-present

Conference Proceedings Citation Index – Science (CPCI-S)--1990-present

Conference Proceedings Citation Index – Social Science & Humanities (CPCI-SSH)--1990-present

Book Citation Index – Science (BKCI-S)--2005-present

Book Citation Index – Social Sciences & Humanities (BKCI-SSH)--2005-present

Emerging Sources Citation Index (ESCI)--2005-present

Current Chemical Reactions (CCR-EXPANDED)--1985-present

Index Chemicus (IC)--1993-present

**Database:** Scopus
**Platform:** Elsevier
**Date Searched:** April 11, 2022

**Database Date Coverage:** 1788–present

**Date Limits:** Publication date: 1990–2022
**Other Limits/Filters:** Language: English & Spanish

| **Set** | **Concept** | **Search Strategy** |
| --- | --- | --- |
| #1 | HPV or Cervical Cancer AND Helminths | Title-Abs-Key( (((HPV OR {cervical cancer*} OR {cervical neoplasm*} OR {cervical neoplasia*} OR {cervical intraepithelial neoplasia*} OR papillomavirus* OR {papilloma virus*} OR {cervix tumour*} OR {cervix tumor*} OR {cervix cancer*} OR {cervix carcinoma*} OR {cervical tumor*} OR {cervical tumour*} OR {uterine cervix tumor} OR {uterine cervix cancer} OR {uterine cervix carcinoma} OR {uterine cervix carcinoma in situ} OR {Wart virus} OR {papillomavirus infection}) AND NOT (spine OR spinal)) OR ((cancer OR cancers OR cancerous OR neoplasm* OR carcinogen*) AND (cervix OR cervical)) ) AND (parasite* OR parasitic OR parasitosis OR helminth* OR ectoparasit* OR schistosom* OR hookworm* OR {hook worm*} OR ancylostomatoid* OR ancylostom* OR bunostomum* OR necator* OR trichuri* OR whipworm* OR {whip worm*} OR bilharzia* OR bilharziosis OR ascari*) ) |
| #2 | Limits Applied | #1 AND ( LIMIT-TO ( LANGUAGE , "English" ) OR LIMIT-TO ( LANGUAGE , "Spanish" ) ) AND ( LIMIT-TO ( PUBYEAR , 2022 ) OR LIMIT-TO ( PUBYEAR , 2021 ) OR LIMIT-TO ( PUBYEAR , 2020 ) OR LIMIT-TO ( PUBYEAR , 2019 ) OR LIMIT-TO ( PUBYEAR , 2018 ) OR LIMIT-TO ( PUBYEAR , 2017 ) OR LIMIT-TO ( PUBYEAR , 2016 ) OR LIMIT-TO ( PUBYEAR , 2015 ) OR LIMIT-TO ( PUBYEAR , 2014 ) OR LIMIT-TO ( PUBYEAR , 2013 ) OR LIMIT-TO ( PUBYEAR , 2012 ) OR LIMIT-TO ( PUBYEAR , 2011 ) OR LIMIT-TO ( PUBYEAR , 2010 ) OR LIMIT-TO ( PUBYEAR , 2009 ) OR LIMIT-TO ( PUBYEAR , 2008 ) OR LIMIT-TO ( PUBYEAR , 2007 ) OR LIMIT-TO ( PUBYEAR , 2006 ) OR LIMIT-TO ( PUBYEAR , 2005 ) OR LIMIT-TO ( PUBYEAR , 2004 ) OR LIMIT-TO ( PUBYEAR , 2003 ) OR LIMIT-TO ( PUBYEAR , 2002 ) OR LIMIT-TO ( PUBYEAR , 2001 ) OR LIMIT-TO ( PUBYEAR , 2000 ) OR LIMIT-TO ( PUBYEAR , 1999 ) OR LIMIT-TO ( PUBYEAR , 1998 ) OR LIMIT-TO ( PUBYEAR , 1997 ) OR LIMIT-TO ( PUBYEAR , 1996 ) OR LIMIT-TO ( PUBYEAR , 1995 ) OR LIMIT-TO ( PUBYEAR , 1994 ) OR LIMIT-TO ( PUBYEAR , 1993 ) OR LIMIT-TO ( PUBYEAR , 1992 ) OR LIMIT-TO ( PUBYEAR , 1991 ) OR LIMIT-TO ( PUBYEAR , 1990 ) ) |

**Notes:** The limits for publication year (1990–2022) and language (English and Spanish) were applied to the main search using the filter available. The keywords were searched in the title, abstract, or keywords fields. Phrases were enclosed in curly brackets (i.e., { }) to force the searching of the exact terms in order presented. No other limits were applied to the searches.

**Database:** Embase
**Platform:** Elsevier
**Date Searched:** April 11, 2025

**Database Date Coverage:** 1947–present
**Date Limits:** Publication date: 1990–2022
**Other Limits/Filters:** Source: Embase; Language: English & Spanish; Unselect the checkboxes under Mapping

| **Set** | **Concept** | **Search Strategy** |
| --- | --- | --- |
| #1 | HPV or Cervical Cancer | (((HPV:ti,ab OR 'human papilloma virus':ti,ab OR 'human papillomavirus':ti,ab OR 'cervical cancer*':ti,ab OR 'cervical neoplasm*':ti,ab OR 'cervical neoplasia*':ti,ab OR 'cervical intraepithelial neoplasia*':ti,ab OR papillomavirus*:ti,ab OR 'papilloma virus*':ti,ab OR 'cervix tumour*':ti,ab OR 'cervix tumor*':ti,ab OR 'cervix cancer*':ti,ab OR 'cervix carcinoma*':ti,ab OR 'cervical tumor*':ti,ab OR 'cervical tumour*':ti,ab OR 'uterine cervix tumor'/exp OR 'uterine cervix cancer'/exp OR 'uterine cervix carcinoma'/exp OR 'uterine cervix carcinoma in situ'/exp OR 'Wart virus'/exp OR 'papillomavirus infection'/exp) NOT (spine:ti,ab OR spinal:ti,ab)) OR ((cancer:ti,ab OR cancers:ti,ab OR cancerous:ti,ab OR neoplasm*:ti,ab OR carcinogen*:ti,ab) AND (cervix:ti,ab OR cervical:ti,ab))) |
| #2 | Helminths | (parasite*:ti,ab OR parasitic:ti,ab OR parasitosis:ti,ab OR helminth*:ti,ab OR ectoparasit*:ti,ab OR schistosom*:ti,ab OR hookworm*:ti,ab OR "hook worm*":ti,ab OR bunostomum*:ti,ab OR necator*:ti,ab OR ascari*:ti,ab OR trichuri*:ti,ab OR whipworm*:ti,ab OR 'whip worm*':ti,ab OR bilharzia*:ti,ab OR bilharziosis:ti,ab OR ancylostomatoid*:ti,ab OR ancylostom*:ti,ab OR helminth*:ti,ab OR 'parasitosis'/de OR 'helminthiasis'/exp OR 'ancylostomiasis'/exp OR 'necatoriasis'/exp OR 'Necator'/exp OR 'Trichuris'/exp OR 'trichuriasis'/exp OR 'hookworm'/exp OR 'Ascaris'/exp OR 'ascariasis'/exp OR 'Ancylostoma'/exp OR 'hookworm infection'/exp OR 'Schistosoma'/exp OR 'schistosomiasis'/exp) |
| #3 |  | #1 AND #2 |
| #4 | Limits Applied | #3 AND [1990-2022]/py AND ([english]/lim OR [spanish]/lim) AND [embase]/lim |

**Notes:** The limits for publication year (1990–2022), source (Embase), and language (English and Spanish) were applied to the main search using the filters available in Embase. The keywords were searched in the title and abstract fields (i.e., :ti,ab), and the EMTREE controlled vocabulary terms were searched as /de or if the EMTREE term was exploded to automatically include all narrower terms this was indicated with /exp. Phrases were enclosed in single quotation marks to force the searching of the exact terms in order presented. No other limits were applied to the searches.

**Database:** Global Health
**Platform:** CABI
**Date Searched:** April 11, 2025

**Database Date Coverage:** 1973–present
**Date Limits:** Publication date: 1990–2022
**Other Limits/Filters:** Language: English & Spanish

| **Set** | **Concept** | **Search Strategy** |
| --- | --- | --- |
| #1 | HPV or Cervical Cancer | Title: (((HPV OR "cervical cancer*" OR "cervical neoplasm*" OR "cervical neoplasia*" OR "cervical intraepithelial neoplasia*" OR papillomavirus* OR "papilloma virus*" OR "cervix tumour*" OR "cervix tumor*" OR "cervix cancer*" OR "cervix carcinoma*" OR "cervical tumor*" OR "cervical tumour*" OR "uterine cervix tumor" OR "uterine cervix cancer" OR "uterine cervix carcinoma" OR "uterine cervix carcinoma in situ" OR "Wart virus" OR "papillomavirus infection") AND NOT (spine OR spinal)) OR ((cancer OR cancers OR cancerous OR neoplasm* OR carcinogen*) AND (cervix OR cervical)) ) |
| #2 | HPV or Cervical Cancer | Abstract: (((HPV OR "cervical cancer*" OR "cervical neoplasm*" OR "cervical neoplasia*" OR "cervical intraepithelial neoplasia*" OR papillomavirus* OR "papilloma virus*" OR "cervix tumour*" OR "cervix tumor*" OR "cervix cancer*" OR "cervix carcinoma*" OR "cervical tumor*" OR "cervical tumour*" OR "uterine cervix tumor" OR "uterine cervix cancer" OR "uterine cervix carcinoma" OR "uterine cervix carcinoma in situ" OR "Wart virus" OR "papillomavirus infection") AND NOT (spine OR spinal)) OR ((cancer OR cancers OR cancerous OR neoplasm* OR carcinogen*) AND (cervix OR cervical)) ) |
| #3 | HPV or Cervical Cancer | Thesaurus Terms: ("human papillomaviruses" OR "cervical cancer" OR "cervical intraepithelial neoplasia" OR "cervical neoplasms") |
| #4 |  | #1 OR #2 OR #3 |
| #5 | Helminths | Title: (parasite* OR parasitic OR parasitosis OR helminth* OR ectoparasit* OR schistosom* OR hookworm* OR "hook worm*" OR ancylostomatoid* OR ancylostom* OR bunostomum* OR necator* OR trichuri* OR whipworm* OR "whip worm*" OR bilharzia* OR bilharziosis OR ascari*) |
| #6 | Helminths | Abstract: (parasite* OR parasitic OR parasitosis OR helminth* OR ectoparasit* OR schistosom* OR hookworm* OR "hook worm*" OR ancylostomatoid* OR ancylostom* OR bunostomum* OR necator* OR trichuri* OR whipworm* OR "whip worm*" OR bilharzia* OR bilharziosis OR ascari*) |
| #7 | Helminths | Thesaurus Terms: ("hookworm" OR "hookworms" OR "helminthoses" OR "whipworm infection" OR "trichuriasis" OR "trichuriasis" OR "Trichuris" OR "ascariasis" OR "Ascaris" OR "schistosomiasis" OR "Ancylostomatidae" OR "Bunostomum" OR "Necator (Nematoda)") |
| #8 |  | #5 OR #6 OR #7 |
| #9 |  | #4 AND #8 |
| #10 | Limits Applied | #9 AND Language: English AND Spanish AND Publication Year: 1990–2022 |

**Notes:** The limits for publication year (1990–2022) and language (English and Spanish) were applied to the main search using the filters available. The keywords were searched in the title and abstract fields, and the controlled vocabulary terms were searched in the Thesaurus field. Phrases were enclosed in single quotation marks to force the searching of the exact terms in order presented. No other limits were applied to the searches.
