## Supplemental Data 1 for "Understanding the association between immune modulating helminths and human papillomavirus or cervical cancer: a scoping review"

**Supplemental Information 3 – Codebook**

Objective: This scoping review looked at epidemiologic studies to understand what is currently known about the association between soil-transmitted helminths, Human papillomavirus prevalence and persistence, cervical cancer progression, and to identify gaps in the literature.

Population: Women of any age with cervical cancer or HPV infection

Exposure: soil-transmitted helminths, (specifically, schistosomes, hookworms, ascaris, Trichuris, and bilharzia) (active or past infection)

Outcome: HPV persistence, prevalence, and progression into cervical neoplasia or cancer

Setting: Lower- and middle-income countries

**Data extraction codebook**

| Data Point | Answer Options |
| --- | --- |
| Title |  |
| Author |  |
| Publication Date |  |
| Country |  |
| Sample size |  |
| Age range of participants |  |
| Setting | Urban; Rural; Hospital; Clinic; Community Based Health Clinic |
| Study Objective |  |
| Study Design | Cohort; Case Control; Follow-up; Observational; Cross Sectional; Prospective; Retrospective; Case Study; Medical Records; Molecular Epi |
| Exposure: Type of Helminth | Schistosomiasis; Hookworm; Trichuris; Ascaris |
| Methods used to measure exposure status |  |
| Outcome | HPV; Cervical Cancer; Both |
| Methods used to measure outcome status |  |
| Methods used to measure how exposure and outcome related to one another/comparison |  |
| Main analysis method |  |
| Primary outcome as related to HPV, cervical cancer, and helminth infection |  |
| Comments raised in manuscript |  |
| Limitations listed in manuscript |  |
| Gaps/Next Steps |  |
